# BLOOD PROTEOMICS AND PAIN - A TRANSLATIONAL STUDY TO PROGNOSTICATE PAIN PHENOTYPES AND ASSESS NEW BIOMARKERS FOR PREVENTING PAIN IN HUMANS

**DOI:** 10.1101/2024.07.04.24309933

**Authors:** Daniel Segelcke, Julia R. Sondermann, Christin Kappert, Bruno Pradier, Dennis Görlich, Manfred Fobker, Jan Vollert, Peter K. Zahn, Manuela Schmidt, Esther M. Pogatzki-Zahn

## Abstract

Personalized strategies in pain management and prevention should be based on individual risk factors as early as possible, but the factors most relevant are not yet known. An innovative approach would be to integrate multi-modal risk factors, including blood proteomics, in predicting high pain responders and using them as targets for personalized treatment options. Here, we determined and mapped multi-modal factors to prognosticate a phenotype with high risk of developing pain and hyperalgesia after an experimental incision in humans. We profiled unbiased blood plasma proteome signature of 26 male volunteers, assessed psychophysical and psychological aspects before incision injury. Outcome measures were pain intensity ratings and the extent of the area of hyperalgesia to mechanical stimuli surrounding the incision as a proxy for central sensitization. Phenotype-based stratification resulted in the identification of low- and high-responders for the two different outcome measures. Logistic regression analysis revealed prognostic potential for blood plasma proteins and for psychophysical and psychological parameters. The combination of certain parameters increased the prognostic accuracy for both outcome measures, exceeding 97%. In high-responders, term-term-interaction network analysis showed a proteome signature of a low-grade inflammation reaction. Intriguingly, *in silico* drug repurposing indicates a high potential for specific antidiabetic and anti-inflammatory drugs already available. In conclusion, we show an integrated pipeline that provides a valuable resource for patient stratification and the identification of (i) multi-feature prognostic models, (ii) treatment targets, and (iii) mechanistic correlates that may be relevant for individualized management of pain and its long-term consequences.

**One Sentence Summary:** Unbiased identification of blood protein signatures in a translational human postoperative pain model provides new targets for managing pain.

## INTRODUCTION

Pain is a major burden of disease and sufficient people suffer lifelong once pain has become chronic (*1*). Although treatment options are available, their efficacy is rather low and heterogeneous from patient to patient. In addition, side effects of several drugs limit their use and have contributed to the opioid epidemic in many countries, especially the US (*2, 3*). A major problem is that chronic pain underlies multiple mechanisms ranging from biological to psychological, mental, societal, among others. Pain and the underlying mechanisms differ among patients with the same disease. Thus, identifying the individual factors responsible for the development of chronic pain in each patient is an important mean relevant for developing novel treatment strategies and personalized approaches to prevent pain best before it has become chronic (*4–6*).

A clinical model very useful for investigating the development of chronic pain is chronic pain after surgery, also termed chronic post-surgical pain (CPSP). CPSP is defined as pain that persists after a surgical procedure beyond the healing process, i.e., for at least three months after surgery (*7*). CPSP is only one example of chronic pain, but with a high prevalence and relevance worldwide. It is estimated that over 300 million surgical procedures are performed worldwide each year, with an estimate of approximately 10% of patients developing CPSP that can limit the patient’s quality of life and represent a considerable socio-economic burden for both the patient and society (*2, 4, 8*). The importance of CPSP was recently highlighted by its inclusion in the International Classification of Diseases catalogue (ICD-11th version, (*7*)). One problem is the knowledge gap related to mechanisms relevant for CPSP. As shown, the development of CPSP is multi-factorial and a complex bio-psycho-social contribution is most likely (*4*). Furthermore, mechanisms of CPSP likely differ between patients and one treatment might not fit all (*4*). All drugs assessed so far for the prevention of CPSP failed (*9*) and prognosis of those patients developing CPSP is as well not yet possible (*10, 11*) one reason for both, failure of prediction and prevention of CPSP, is that biological factors are not integrated in such approaches (*12*). For instance, none of the prognostic prediction models for CPSP so far have used unbiased proteomic analysis (*11*); an unbiased proteomic approach might not only provide additional power to the risk assessment but would also show new targets for new treatment approaches.

Blood represents an easily obtainable diagnostic material in routine clinical practice. Blood plasma, with its diverse array of protein categories, including classical plasma proteins, tissue leakage proteins, and particularly signaling proteins such as hormones, growth factors, and cytokines, serves as a rich resource for modern biomarker discovery (*13*). The presence and levels of these proteins can reflect the body’s physiological and pathological states, making plasma an accessible window into overall health and disease mechanisms. Analyzing blood plasma enables the identification of biomarkers for diagnosing diseases, monitoring health conditions, guiding therapeutic interventions, or prognosticating outcomes (*14*), underscoring its critical role in medical research and personalized medicine. Recent technological advances in sample collection and processing have made it possible for several hundred proteins to be detectable in one sample. However, because of the wide dynamic range and fluctuating abundances of numerous proteins, plasma proteomics remain challenging (15). Clinical laboratories often use highly optimized single-protein assays, allowing for a narrow/diagnostic snapshot while neglecting the complexity of (patho-) physiological conditions. In contrast, mass spectrometry (MS) offers great potential for rapidly and robustly measuring several hundred proteins in a single human sample. Although MS-systems have not yet found their way into clinical routine guidelines, their versatile workflows, which allow for targeted as well as unbiased approaches, make them attractive for many clinical applications and research questions. So far, MS-based proteomics has been used in clinical settings in oncology, endocrinology, and rheumatology, supporting medical decision tools and providing more profound knowledge of underlying molecular mechanisms (*15–21*).

We hypothesize that blood plasma proteome analysis will enable identification of biological predictors associated with and likely relevant for the development of different phenotypes after an experimental incision - a model of pain (*22, 23*). Furthermore, integrating psychophysical and psychosocial parameters together with results from the plasma proteome analysis might provide novel tools for the prognosis of chronic pain after surgery and beyond. Our multi-level analysis enabled us to determine several prognostic factors, which we used to develop prognostic prediction models for different pain-related outcomes, like those associated with high pain and/or a large area of secondary hyperalgesia (a proxy for central sensitization and a risk factor of CPSP (*24–26*), after experimental incision injury. Finally, by using an *in silico* drug repositioning approach, we identified drugs which may modulate protein-protein-interaction (PPI) networks with high pain responders as potential drugs to prevent chronic pain.

## RESULTS

### Pre-incisional blood plasma proteome, sensory phenotyping and psychological volunteer characterization

A cohort of 26 male volunteers, with a mean age of 23.9 years (SD, ±3.64), was enrolled in the study with specific in- and exclusion criteria (Fig. 1A, Table S1). Previously, we have employed this cohort to examine the proteome profile of the skin following an incision injury and directly compare this profile to a comparable injury in mice (*27*). Prior to performing the incision injury (Fig. 1B), multimodal phenotyping of the volunteers (Fig. 1C) was performed at the baseline (BL). Incision pain (IncP) in a one-hour period and secondary hyperalgesia area (HA1) one-hour post-incision were assessed as forms of peripheral and central sensitization, respectively, (Fig. 1C). The prognostic prediction models were developed from multi-modal BL-datasets, blood plasma proteome, sensory phenotyping, and psychological profile, and to predict both outcome measures post-incision (Fig. 1D).

**Fig. 1.**
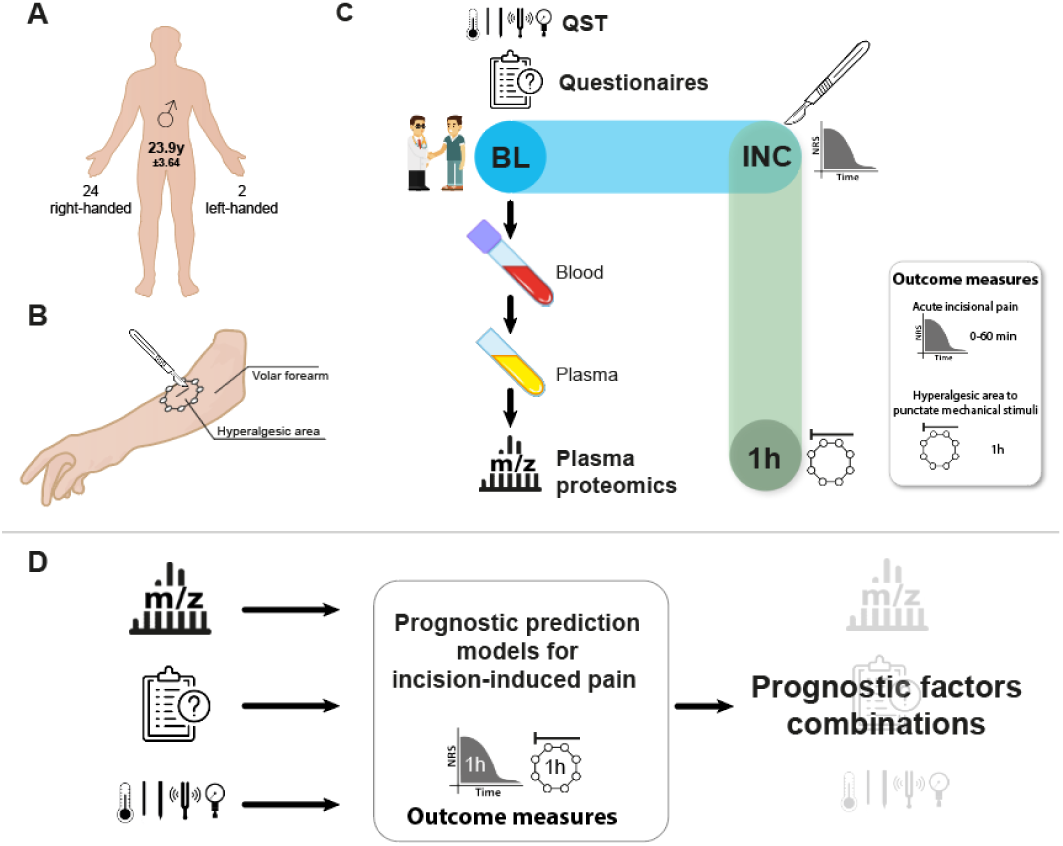
Study design and methodology for predicting incisional outcome measures. **(A)** A total of 26 male volunteers (24 right-handed) with an average age of 23.9 years (y ± SD) were enrolled in the study. **(B)** Experimental incision (INC) was performed with a scalpel on the volar lower am (side randomized). The skin incision had 4 mm length and 7 mm depth. The secondary hyperalgesic area to punctate mechanical stimuli around the incisional injury was determined 1 h post incision. **(C)** Twenty-four h before incision (baseline, BL), the volunteer was medically briefed, complete qualitative sensory testing (QST) was performed in the incision area, five different questionnaires were completed, and blood was collected. Quantitative mass spectrometry was used to identify blood plasma proteome. **(D)** Analysis of BL’s datasets allowed determination of the prognostic potential of single and combinatorial factors to predict pain phenotype, here, incisional pain graded on a numeric rating scale (NRS) and the dimension of the hyperalgesic area around the injury, after incision.

The blood plasma proteome (Fig. 2A-C), the sensory phenotype (Fig. 2D, Table S2), and the psychological profile (Fig. 2E-H, Table S3) of the participants were assessed prior to incision injury (baseline, BL). All volunteers exhibited a typical sensory phenotype (Figure 2D, Table S2) and did not manifest any significant psychosocial outcomes (Figure 2E-H, Table S3). Due to inadequately high-lipid concentration in the blood plasma, one volunteer was excluded from all subsequent analyzes. Considering our quality and cut-off criteria (please see methods for details), we identified and quantified 383 protein groups (hereafter coined “proteins”) across samples (Fig. 2A, Fig. S1). In accordance to the Human Protein Atlas (https://www.proteinatlas.org/humanproteome/), identified proteins could be categorized into *blood plasma proteins*, *tissue leakage proteins* showing no clear function in blood and *signal proteins,* like cytokines or hormones usually found in the low concentration range. To gain more insights into the function of all quantified proteins, we performed a Gene Ontology (GO) enrichment analysis and REACTOME pathway analysis using the STRING web interface (please see methods for additional details; see details in Fig. S1). As expected, a significant portion of the top results were associated with their role in oxygen and nutrient transportation, immune response, and blood clotting (see Fig. S1).

**Fig. 2.**
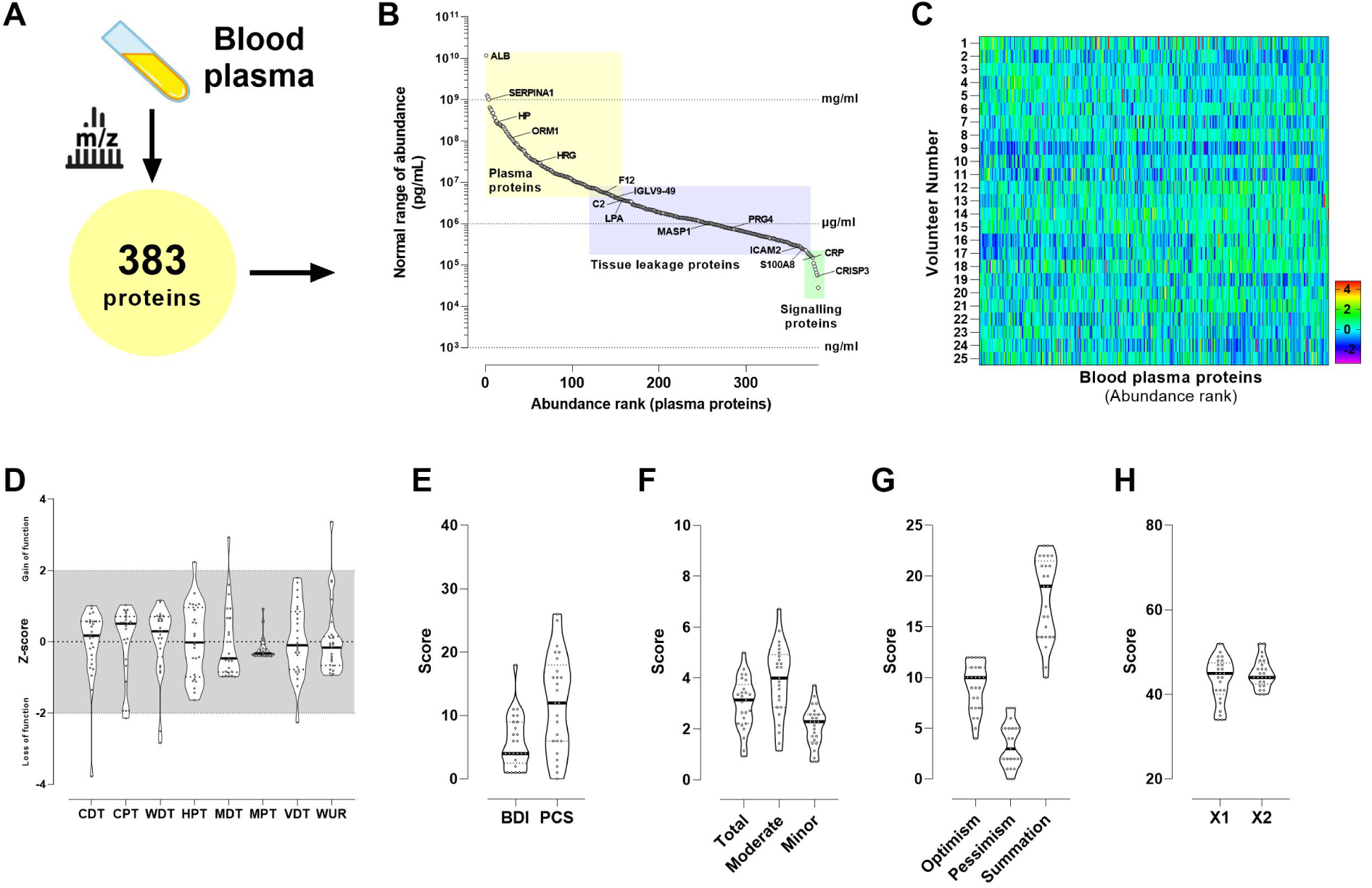
Unbiased discovery proteome profiling of blood plasma proteins, psychophysical, and psychosocial phenotyping before incision injury in male volunteers. **(A)** Identification of 383 pre-incisional blood plasma proteins. When comparing quality marker panels for blood plasma samples, 58 assumed contaminations were found in the identified and quantified protein groups. **(B)** UniProtKB keyword annotations and their enrichment across the protein abundance spectrum, categorized into “blood plasma proteins,” “tissue leakage proteins,” and “signaling proteins” as per The Human Protein Atlas (https://www.proteinatlas.org/humanproteome/blood+protein/proteins+detected+in+ms). Exemplary proteins contributing to keywords are highlighted in red. ALB, albumin; SERPINA1, serpin family A member 1; HP, haptoglobin, ORM1, alpha-1-acid glycoprotein 1; HRG, histidine-rich glycoprotein; KLKB1, plasma kallikrein; F12, coagulation factor XII; C2, complement C2; IGLV9-49, Immunoglobulin Lambda Variable 9-49; MASP1, Mannan-binding lectin serine protease 1; F7, coagulation factor VII; S100A8, S100 calcium binding protein A8; ICAM2, Intercellular adhesion molecule 2; CRISP3, Cysteine Rich Secretory Protein 3. **(C)** The heatmaps show z-weighted median intensities of the 383 identified plasma proteins for each of the 25 volunteers. **(D)** Detection of pre-incisional somatosensory perception to natural stimuli (thermal and mechanical) was achieved by a complete quantitative sensory testing (QST) battery on both forearms. Raw QST-data were transformed to a standard normal distribution (Z-transformation) to achieve QST parameters independent of their physical dimension. The contralateral forearm served as the control side. Z-scores above ‘0’ show a gain of function (more sensitive) and below a loss of function (less sensitive). Values above or below two-fold standard deviation (SD, dotted line) show pathological QST-scores, (Violin Plots, Median ± 95% CI). Abbreviations: CDT, cold detection threshold; CPT, cold pain threshold; HPT, heat pain threshold; MDT, mechanical detection threshold; MPS, mechanical pain sensitivity; MPT, mechanical pain threshold; PPT, pressure pain threshold; TSL, thermal sensory limen; VDT, vibration detection threshold; WDT, warmth detection threshold; WUR, wind-up ratio. **(E)** The individual distribution of the Beck Depression Inventory (BDI) and Pain Catastrophizing Scale (PCS) scores are shown, which illustrate the psychological profiles of the study participants regarding depression and pain catastrophizing, (Violin Plots, Median ± 95% CI). **(F)** The individual distribution of the Perceived Stress Questionnaire (PSQ-Total, Moderate, and Minor) scores are shown, which illustrate the psychological profiles of the study participants regarding stress perception, (Violin Plots, Median ± 95% CI). **(G)** Individual scores of the Life Orientation Test (LOT-Optimism, LOT-Pessimism, and LOT Summation) scores, illustrating the psychological profiles of the study participants in terms of optimism and pessimism, (Violin Plots, Median ± 95% CI). (H) Individual distribution of the State-Trait Anxiety Inventory (STAI x1 and STAI x2) scores, illustrating the psychological profiles of the study participants in terms of anxiety levels, (Violin Plots, Median ± 95% CI).

### Developing a pipeline for prediction models for post-incisional outcome measures based on pre-incisional multi-modal datasets

Following the incision injury on the volar forearm (Fig. 1B), the volunteers experienced acute pain at the site of incision (Fig. S2A), demonstrated heightened sensitivity to punctate mechanical stimuli at the incision site (Fig. S2B), and developed hyperalgesia to punctate mechanical stimuli (Fig. S2C). Ultimately, we designated IncP and HA1 as outcome measures for incision-induced peripheral and central sensitization to evaluate the prognostic capabilities of proteomic, psychophysics, and psychological analyzes as single markers and multi-modal prognostic prediction signatures for incision injury (Fig. 1D). Stratification of the volunteers resulted in 9 individuals classified as high-responders (with a range of 36.1 to 458.5) and 16 individuals classified as low-responders (with a range of 1.7 to 30) for the IncP outcome measure (Fig. 3A). By evaluating individual pre-incisional multi-modal features (Fig. 2) and incorporating them, we created prognostic predictive models for outcome measures. Prognostic proteome signatures were ranked and selected using logistic regression analysis, revealing the best performing classifiers (26). Receiver operating characteristic (ROC) curves and the corresponding area under the curve (ROC_AUC_) determined classifiers for either high-(ROC_AUC_ ↑) or low-responders (ROC_AUC_ ↓) for proteome (Fig 3B), psychophysical parameters (Fig 3C), and multiple-construct psychological profile (Fig 3D). To sum up, among the 311 blood plasma proteins (reduction of the total protein number by 72 possible blood contaminants (*28*)) that were identified, 70 proteins displayed an ROC_AUC_ > 0.6, while 72 proteins showed an ROC_AUC_ < −0.6 (Fig. 3B) for IncP. The Cold Pain Threshold (CPT) and Wind-Up-Ratio (WUR) were determined to be classifiers for high-responders, while the Mechanical Pressure Threshold (MPT) was observed to be linked with low-responders (Fig. 3C). Stress, as measured by the Perceived Stress Questionnaire (PSQ) in both total and moderate forms, was used as a classifier to identify high-responders (elevated) among psychological metrics (Fig. 3D). On the other hand, the evaluation of dispositional optimism at the individual level, as measured by the Life Orientation Test (LOT), served as a classifier for individuals with low responses (↓) (Fig. 3D). Following an analysis of the outcome measure HA1, it was determined that 12 participants could be classified as high-responders, displaying a HA range of 63.5 to 184 cm^2^. Furthermore, 13 individuals were identified as low-responders, exhibiting a range of 1 to 63.5 cm^2^ (Figure 3E). Interestingly, on the proteome level, 60 proteins exhibited an ROC_AUC_ value greater than 0.6, showing a stronger predictive accuracy for HA1. Additionally, 55 proteins were characterized by an ROC_AUC_ value lower than −0.6 (Fig. 3F). The prognostic parameters for individuals classified as high-responders were determined to be the Warm Detection Threshold (WDT), Heat Pain Threshold (HPT), Mechanical Detection Threshold (MDT), Vibration Detection Threshold (VDT), and WUR (Fig 3G). The psychological measures for depression (Beck Depression Inventory - BDI), pain catastrophizing (Pain Catastrophizing Scale - PCS), life orientation (Life Orientation Test - LOT), and minor stress (Perceived Stress Questionnaire - Minor - PSQ-Minor) served as indicators for identifying individuals classified as High Responders for HA1 (Fig. 3H).

**Fig. 3.**
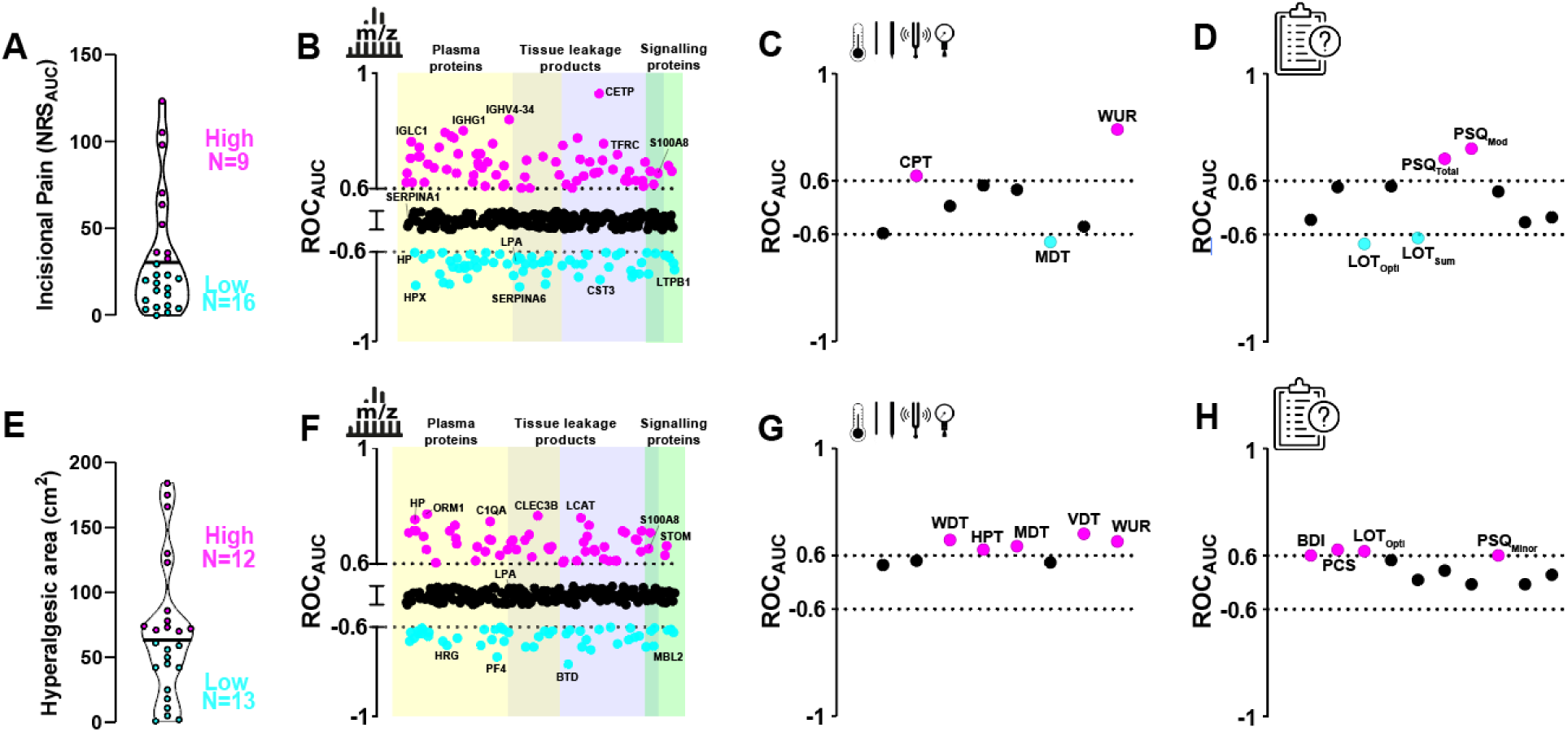
Prognostic value of pre-incisional blood plasma proteome profile, psychophysics, and psychosocial phenotype for post-incisional outcome measures. **(A)** Volunteers were asked to judge the ongoing pain caused by the incision using the Numeric Rating Scale (NRS; 1-100). Integrated data from the first hour after the incision (first every minute for ten minutes, then every 5 minutes for 1h) were analyzed, and the areas under the curve (AUC) were plotted. Twenty-five volunteers (one volunteer was excluded because of technical reasons) were stratified into low (below the mean; N=16; cyan) or high (above the mean, N=9; magenta) responders depending on the rated pain perception within the acute post-incisional phase (1 h post-injury). **(B)** Prognostic value of each of 311 pre-incisional blood plasma proteins for high (magenta, AUC>0.6, 70 proteins) and low-responders (cyan, AUC<0.4, 72 proteins) for incisional pain. **(C)** Prognostic value of all parameters of psychophysical testing for high (magenta, AUC>0.6, CPT and WUR) and low-responders (cyan, AUC<0.4, MDT) for incisional pain. **(D)** Prognostic value of psychosocial questionnaires for high (magenta, AUC>0.6, PSQ-Total, PSQ-Moderate) and low-responders (cyan, AUC<0.4, LOT-Optimism, LOT-Summation) for incisional pain. **(E)** Upon incision injury, hyperalgesic area (HA) was assessed post 1 h (HA1) in 25 volunteers. Phenotyping was achieved by determining the total cohort’s mean HA (68.65 cm^2^). Volunteers with a lower HA1 were categorized as “low-responders” (N=13), and those with higher mean as “high-responders” (N=12). **(F)** Prognostic value of each of 311 pre-incisional blood plasma proteins for high (magenta AUC>0.6, 60 proteins) and low-responders (cyan, AUC<0.4, 55 proteins) for hyperalgesic area 1h post-incision (HA1). **(G)** Prognostic value of all parameters of psychophysical testing for high (magenta, AUC>0.6, WDT, HPT, MDT, VDT, WUR) for HA1. **(H)** Prognostic value of psychosocial questionnaires for high (magenta, AUC>0.6, BDI, PCS, LOT-Optimism, and PSQ-Minor) for HA1. Abbreviations: CDT, cold detection threshold; CPT, cold pain threshold; HPT, heat pain threshold; MDT, mechanical detection threshold; MPS, mechanical pain sensitivity; MPT, mechanical pain threshold; PPT, pressure pain threshold; TSL, thermal sensory limen; VDT, vibration detection threshold; WDT, warmth detection threshold; WUR, wind-up ratio; BDI, Beck Depression Inventory; PCS, Pain Catastrophizing Scale; PSQ-Total, Perceived Stress Questionnaire-Total; PSQ-Moderate, Perceived Stress Questionnaire-Moderate; PSQ-Minor, Perceived Stress Questionnaire-Minor; LOT-Optimism, Life Orientation Test-Optimism; LOT-Pessimism, Life Orientation Test-Pessimism; LOT Summation, Life Orientation Test Summation; STAI x1, State-Trait Anxiety Inventory Form X1; STAI x2, State-Trait Anxiety Inventory Form X2.

The utilization of data-driven combinations involving two proteins resulted in improved accuracy for both outcome measures, with a mean value of 0.81 [0.67-0.99, ΔAUC=0.13] for IncP (Fig. 4A) and 0.85 [0.7-0.96, ΔAUC=0.13] for HA1 (Fig. 4B), when compared to the utilization of only one protein (IncP, 0.68 [0.61-0.93]). Furthermore, the integration of three plasma proteins resulted in an increased accuracy, reaching an average of 0.86 [0.67-1] for IncP and 0.85 [0.7-1] minimally (ΔAUC=0.05). Data-driven combination between a protein and psychological factor or a protein and psychophysical factor did not increase the accuracy for both outcome measures compared to the individual factors. The combination of all three dimensions resulted in a framework that views post-incisional outcomes from a molecular, psychological and sensory perspective, reflecting the complex nature of underlying processes (Fig. 4). On average, an accuracy of 0.76 [0.32-0.97] and 0.74 [0.34-0.96] was achieved for high-responders for IncP and HA1, respectively. The TOP10 combinations for both outcome measures differed in their composition, while the accuracy of the prediction was comparable.

**Fig. 4.**
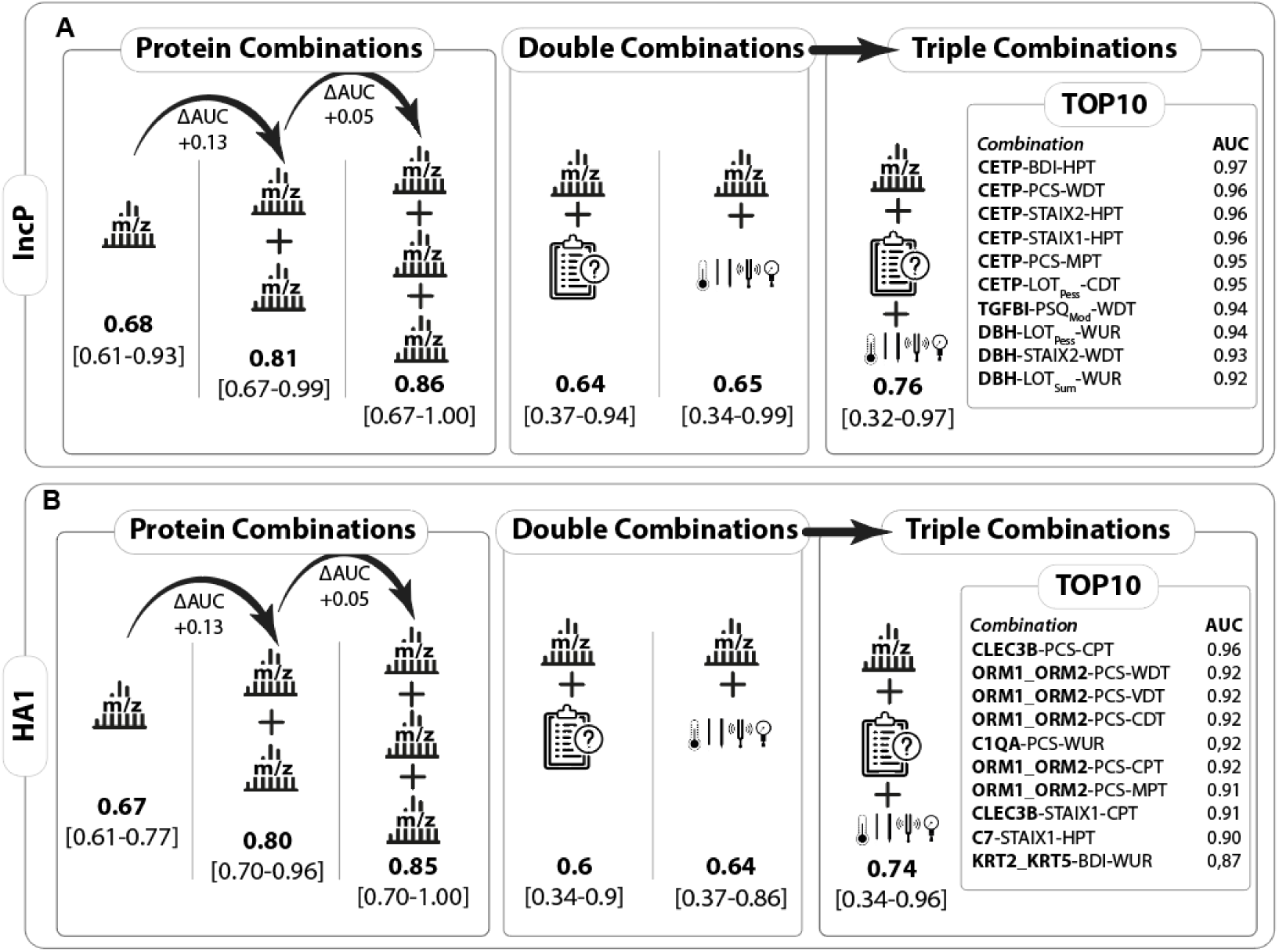
Improved accuracy in predicting post-incision outcomes using data-driven combinations. **(A)** For IncP, the use of two proteins significantly improved accuracy compared to a single protein. Integrating three proteins further increased accuracy. Combining a protein with psychological or psychophysical factors did not enhance accuracy. A framework combining molecular (bold font), psychological, and sensory perspectives showed good accuracy for high-responders. **(B)** For HA1, the combination of two proteins improved accuracy, which remained stable with three proteins. Combining a protein (bold font) with psychological or psychophysical factors did not improve accuracy. The comprehensive approach also yielded good accuracy for high-responders. The TOP10 combinations for both outcomes varied in composition, but prediction accuracy was comparable, highlighting the importance of integrating multiple dimensions to understand post-incision responses. Aberrations: CETP=Cholesteryl Ester Transfer Protein, CGT=Cystathionine Gamma-Lyase, C1QA=Complement C1qa, CLEC3B=C-Type Lectin Domain Family 3 Member B, DBH=Dopamine Beta-Hydroxylase, HPT=Heat Pain Threshold, KRT2=Keratin 2, KRT5=Keratin 5, ORM1=Orosomucoid 1, ORM2=Orosomucoid 2, PCP=Pyridoxal Phosphate, TGFBI=Transforming Growth Factor Beta Induced, BDI=Beck Depression Inventory, PCS=Pain Catastrophizing Scale, PSQMOD=Perceived Stress Questionnaire-Moderate, STAI x1=State-Trait Anxiety Inventory Form X1, STAI x2=State-Trait Anxiety Inventory Form X2, CDT=Cold Detection Threshold, CPT=Cold Pain Threshold, MPT=Mechanical Pain Threshold, VDT=Vibration Detection Threshold, WDT=Warmth Detection Threshold, WUR=Wind-Up Ratio. Mean ROC_AUC_ [min-max Range].

### Molecular insights into peripheral and central Sensitization post-incision: prognostic models and network-based drug repositioning

Next, we aimed at exploring molecular differences in pre-incisional proteome signatures, which may hint towards differential mechanisms for developing high- and low-responder phenotypes in terms of post-incisional outcome (pain intensity (IncP) and hyperalgesia (HA1) as a proxy of central sensitization). To this end we employed network-based drug repositioning analysis (*25*), which seeks to leverage existing therapeutic drugs to modulate disease-associated signaling networks. Comparing low-(Fig. 5A) and high-(Fig 5B) responder categories for both outcome variables, we observed distinct and overlapping protein signatures. (Fig. 5A, B). Enriched term-term-interaction (TTI) network analysis was performed for IncP (Fig. 5C) and HA1 (Fig. 5D), uncovering terms that were enriched in high-responders for both outcomes, with a particular emphasis on clusters related to the innate immune and complement system. The identification of pre-incisional classifier proteins in the cascades of the complement system in high-responders for both outcomes could potentially reflect a pre-incisional low-grade inflammatory condition.

**Fig. 5.**
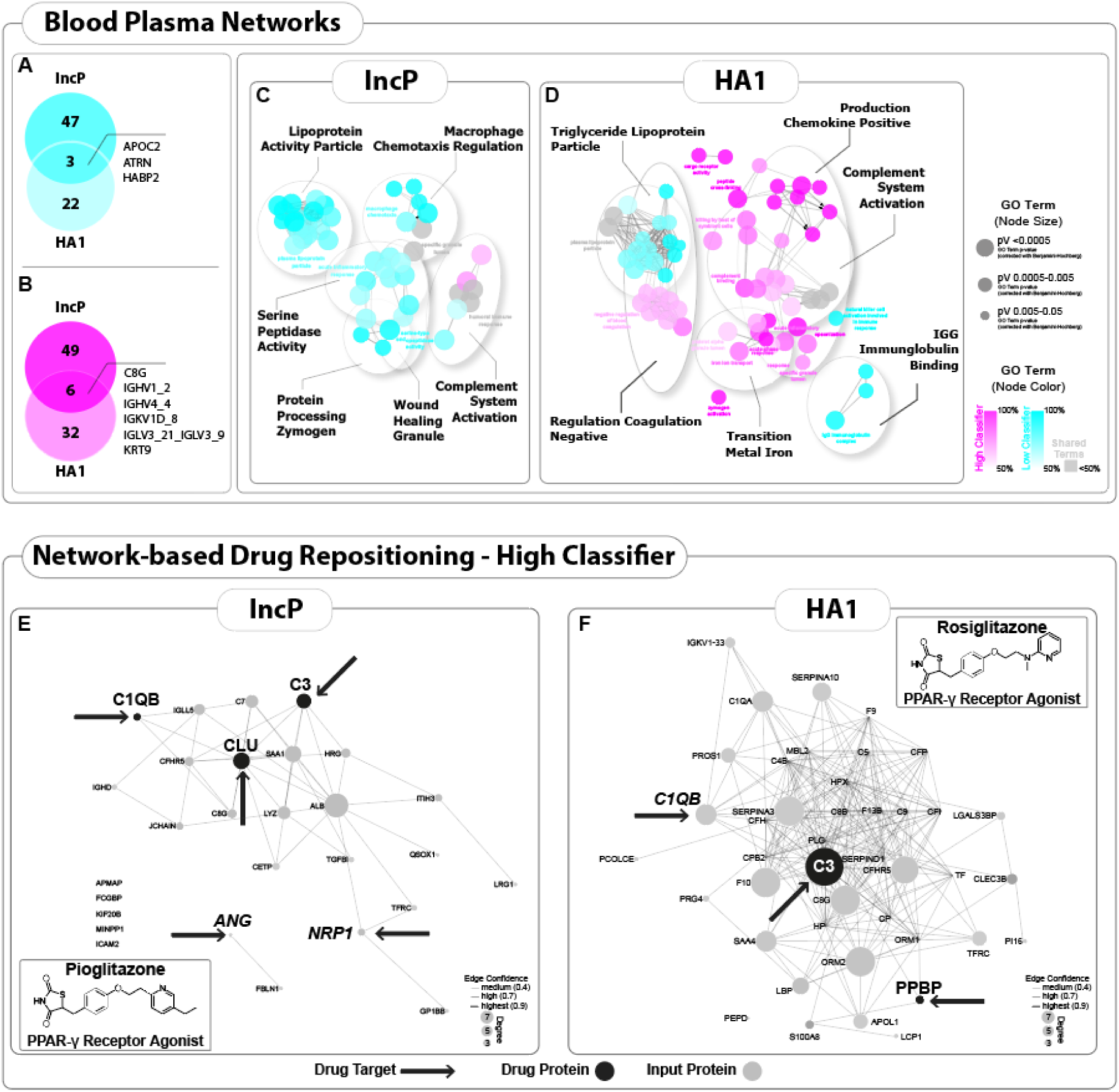
Molecular mechanistic understanding and network-based drug repositioning analysis for pre-incisional biomarker signatures. **(A)** Pre-incisional protein classifier patterns for low-responders. A total of 47 unique proteins were identified for IncP and 22 for HA1 in low-responders, with an overlap of 3 proteins: APOC2, ATRN, and HABP2. **(B)** Pre-incisional protein classifier patterns for high-responders. A total of 49 unique proteins were identified for IncP and 32 for HA1 in high-responders, with an overlap of 6 proteins, specifically C8G, KRTP, and 7 different immunoglobulins. **(C, D)** The functional grouping of the enriched term-term interaction (TTI) network analysis, along with its associated proteins, was performed using ClueGO (v2.5.8), resulting in functional clusters (AutoAnnotate 1.3). **(C)** TTI-network analysis for IncP, highlighting terms exclusively present in high-responders, particularly clusters related to the innate immune system and the complement system via the classical and alternative pathways. (D) TTI-network analysis for HA1, emphasizing terms exclusively present in high-responders, with a focus on the complement system and innate immune response pathways. The data reveals significantly enriched GO terms, as indicated by P-values below 0.05, after applying the Benjamini-Hochberg correction. These terms are observed within a functionally grouped network, which accurately represents their interconnected relationships. Each node corresponds to a molecular function or process. Node colors are associated with phenotyping (high-responder in magenta shades; low-responder in cyan shades). Gray shades reflect unspecific terms which belong to both responder types. **(E)** Protein-protein-interaction (PPI) network drug repositioning results for IncP, showing the presence of the PPARγ agonist Pioglitazone and its associated drug protein proteins (CLU, C3, C1QB), including ANG and NRP1 as input proteins. **(F)** PPI-network drug repositioning results for HA1, indicating the PPARγ agonist Rosiglitazone and its associated drug proteins (C3, PPBP), with C1QB also identified as an input protein.

**Table 1.** Network-based Drug Repositioning with PharmOmics: Detailed Z-Scores, Rankings, and P-Values.

| Drug | Tissue | Z_score | Rank | P-value |
| --- | --- | --- | --- | --- |
| Atorvastatin | hematopoietic system | -3.59 | 0.9968 | 0.000 |
| Immunosuppressant | immune system | -2.55 | 0.9959 | 0.005 |
| Alkylating agent | immune system | -2.55 | 0.9951 | 0.005 |
| Rosiglitazone | hematopoietic system | -2.51 | 0.9951 | 0.006 |
| Infliximab | hematopoietic system | -2.50 | 0.9943 | 0.006 |
| Cyclophosphamide | immune system | -2.48 | 0.9935 | 0.007 |
| Anti-TNF-alpha antibody | hematopoietic system | -2.47 | 0.9927 | 0.007 |
| Rosuvastatin | hematopoietic system | -2.46 | 0.9967 | 0.007 |
| Antimetabolite | immune system | -2.39 | 0.9959 | 0.009 |
| Arsenic trioxide | hematopoietic system | -2.31 | 0.9783 | 0.010 |
| Fludarabine | immune system | -2.27 | 0.9934 | 0.012 |
| COX inhibitor | hematopoietic system | -2.27 | 0.9759 | 0.012 |
| Antidiabetic | hematopoietic system | -2.24 | 0.9918 | 0.013 |
| PPARγ gamma agonist | hematopoietic system | -2.21 | 0.9910 | 0.013 |
| Etoposide | immune system | -2.17 | 0.9780 | 0.015 |

Network drug repositioning was conducted based on responder-type proteome signatures (Fig. 5A, B) using the species- and tissue-specific drug signature platform PharmOmics (http://mergeomics.research.idre.ucla.edu/runpharmomics.php). Upon filtering the results relevant for the hemopoietic and immune system, we obtained 44 significant hits for high-responders and 46 for low-responders. An overlap of specific drug compounds, such as the Anti-CD52 antibody, and various drug classes, including immunosuppressants and statins, was observed among both high- and low-responders. Remarkably, however, “antidiabetic” drugs, specifically, the Peroxisome proliferator-activated receptor gamma (PPARγ) agonists, Pioglitazone and Rosiglitazone, were associated with proteome signatures in high-responders, while Pioglitazone was detected in IncP (Fig. 5E), and Rosiglitazone in HA1 (Fig. 5F). The PPI-network analysis showed the presence of five drug proteins for Pioglitazone in IncP and three proteins as targets for Rosiglitazone in HA1.

## DISCUSSION

This study integrated blood plasma proteomics, psychophysical, and psychological profiles to develop data-driven prognostic prediction models for distinct outcome measurements relevant for pain, e.g. pain intensity (IncP), and mechanical hyperalgesia (HA1), a proxy of central sensitization (*29, 30*) and predictor of chronic pain after surgery (*24–26*). Applying data-driven multi-feature combinations resulted in an enhanced accuracy of prediction models, effectively capturing bio-psycho-social aspects of pain. Protein network analysis revealed a pre-incisional low-grade inflammatory response in high-responders, potentially contributing to heightened IncP and pronounced HA1. Network drug repositioning highlighted PPARγ agonists as possible treatment options for low-grade inflammation in high-responders. Taken together, our approach towards developing a data-driven prognostic prediction model paired with network-based drug repositioning has high utility for future clinical studies using large cohorts to understand and predict both acute and CPSP.

Following a surgical procedure, different outcomes manifest, indicating both peripheral and central sensitization, thereby exerting a substantial impact on the acute and potentially long-term consequences (*4, 31*). Timely identification of high-risk patients is imperative and poses a significant challenge, paired with the necessity of developing novel prognostic prediction models. While several models for the prediction of CPSP have been developed (*10, 11*), they often exhibit limitations that hinder their clinical application. One major limitation is the incomplete understanding of the underlying mechanisms relevant for developing CPSP (*12*). Most models rely on demographics (age or gender), or preoperative pain, while incorporating molecular or genetic markers or psychological factors remains limited (*10, 11*). In fact, no previous study has employed unbiased blood proteome profiling, neither in an experimental or clinical setting, to consider protein. In our prognostic prediction models, integrating plasma proteome signatures acquired prior to incision injury is used to fill this gap in a translational approach (*12*). Blood plasma exhibits several advantages for clinical proteome profiling given that it is easy-to-be-collected (following standardized procedures) and small volumes suffice for downstream molecular analysis(*13, 28, 32*).

We demonstrated that our pipeline effectively identified high- and low-responders for outcomes measures of incision injury. The prognostic prediction models revealed that a combination of two proteins resulted in increased accuracy, while three only added slight improvements reaching a steady state. Despite our efforts to enhance model accuracy by combining protein data with psychological or psychophysical factors, this approach did not yield better performance compared to using individual factors alone. One explanation for this outcome is the inherent complexity and variability in how different types of data interact. Combining proteins, psychological measures, and psychophysical parameters for pain mechanisms may create noise, especially with a limited sample size, as in our translational study. Scale and nature differences of data types could be another factor. Proteomic data is (semi-) quantitative and high-dimensional, while psychological and psychophysical data is qualitative and somehow subjective. Disparities complicate integration, hindering cohesive modeling of diverse factors. Adding more variables increases the risk of model overfitting without adding predictive power. The added complexity might lead to models that perform well on training data but fail to generalize to new, unseen data. Consequently, future studies need to (i) increase sample size, (ii) use independent datasets for validation, (iii) perform longitudinal monitoring, and (iv) incorporate additionally relevant data types, such as multi-omics or imaging data (*33*). Certainly, the success of introducing such pipelines into clinical routines also needs to consider time and economic factors while providing high-throughput and standardized features. Optimizing clinical usage requires considering time, cost, and economic factors (*34*). This involves developing streamlined protocols that balance patient/provider burden with model predictive power. Practicality and scalability of predictive models in clinical settings can be improved through efficient and cost-effective methods.

By analyzing pre-incisional TTI-networks, we sought to understand the molecular differences between high- and low-responders regarding IncP and HA1 outcomes. Proteomic features of responder types pointed towards shared and also distinct pathways measures, suggesting that different biological mechanisms might be at play in low-versus high-responders, influencing their sensitivity to post-surgical pain. The TTI-network analysis highlighted the involvement of innate immune system in high-responders for both outcome measures. These findings are in accordance with known features of persistent pain in various pain etiologies (*35*). The immune-related protein signature in high-responders may show a low-grade inflammatory state. This may facilitate increased reactions to painful injuries or diseases (*36*). Low-grade inflammation refers to a mild state of inflammation representing chronic, yet subtle activation of the immune system. Unlike acute inflammation, chronic low-grade inflammation is typically asymptomatic (*37*). Chronic stress, an unhealthy diet, obesity, lack of exercise, smoking, chronic infections, and autoimmune disorders can all contribute to low-grade inflammation (*38, 39*). Ongoing interactions between the immune system and nervous system can exacerbate and sustain nociceptive signals and be accompanied by the release of pro-inflammatory cytokines and other mediators. While only exploratory, our results suggest that a viable treatment concept in perioperative pain management could involve targeting the low-grade inflammation as a potential preventive strategy for acute and chronic outcomes post-surgery. Here, we employed a computational network-based approach to repurpose drugs and identified PPARγ agonists such as Rosiglitazone and Pioglitazone as potential modulators of low-grade inflammation given their anti-inflammatory capacity (*40*). Indeed, local Rosiglitazone administration reduced hypersensitivity in a mouse incision pain model, dampening post-incisional inflammation and hypersensitivity by modulating macrophage polarity (*41*). Also, the PPARγ agonist Pioglitazone has been reported in several preclinical studies to exert an anti-hypersensitivity effect in a variety of pain models ranging from inflammatory, post-incisional, to neuropathic pain (*42–45*). Considering evidence in preclinical studies for PPARγ agonists in attenuating pain, further studies to investigate the underlying mechanisms and the site of action are warranted (*45*). While a clear advantage of drug repurposing is that the safety profile characteristics of the compound in question are already known, the effective dosage and time point of intervention would need additional investigations. Also, large clinical data sets may optimize the use of network-based models to predict combined effects (both therapeutic and adverse effects) of multiple drugs and their interactions. Such an approach would foster the identification of synergistic drug combinations that target multiple signaling pathways within disease-associated networks (*46–48*). No synergistic drug combinations were found in our setting. Nonetheless it is conceivable that anti-inflammatory drugs, immunomodulators, and metabolic modulators may be combined to synergistically target inflammatory pathways and, by only reduced doses, side effects of drugs are reduced as well.

Taken together, our integrated study provides a stepping-stone towards the development of clinically applicable tools based on bio-psycho-physiological data with great potential for individualized management of post-surgical pain.

## MATERIALS AND METHODS

### Study design

Upon recruitment, by notice boards at the university and medical campus Muenster, 26 male volunteers (age mean 23.9 ± SEM 3.6 years) gave written consent to be part of an incision-induced pain project after being informed about the study and experimental procedure. All volunteers passed the listed in- and exclusion criteria in supplementary table 1. Volunteers in this study were obliged to have an identical breakfast three days in a row. All human experiments were approved by the University Hospital Muenster of the local Ethics Committee of the Medical Faculty (registration no 2018-081-b-S), registered in German Clinical Trials Registry (DRKS-ID: DRKS00016641), and in accordance with the latest version of the Declaration of Helsinki. Data from skin biopsies performed 24 hours after incision from these volunteers are included in a previous publication (*27*).

Comprehensive quantitative sensory testing, baseline psychosocial characterization, and blood sampling were performed before induction of incisional injury. An experimental incision was then made, and the incisional pain and hyperalgesic area were determined at fixed time points.

### Chemicals and Reagents

Chemicals for an in-solution digest of plasma proteins were either purchased as a powder (ABC, IAA, SDC) or in liquid form (DTT, TCEP) from Merck Millipore/Sigma Aldrich (Germany). Protease for enzymatic digestion was a mixture of Trypsin/LysC, Mass Spec grade (Promega, USA). Ethyl acetate for surfactant removal was purchased from Merck Millipore/Supelco in hypergrade purity (Lichrosolv). Acids for organic LC solvents or peptide clean-up were either purchased from Merck Millipore/Sigma Aldrich, Germany (TFA) or Thermo Scientifc, USA (FA). All MS-related solvents (water, ACN) were of hypergrade purity (Lichrosolv) and purchased at Merck Millipore (Supelco). C18 MicroSpin columns (6-60 µg capacity) were supplied by The Nest Group (USA).

### Psychophysical baseline characterization by quantitative sensory testing

Before incision, a comprehensive battery of quantitative sensory testing (QST) was performed on both forearms on the volar aspect (one forearm represents the test area where the incision will later be made, the other forearm represents the control side) to assess the perception of non-painful and painful stimuli of different modalities established by the German Research Network in Neuropathic Pain (DFNS) (*49*). Testing is a subjective (psychophysical) method requiring testing of person cooperation. Briefly, thermal testing will be performed by using a TSA-II NeuroSensory Analyzer (MEDOC, Ramat Yishai, Israel). With this device, thermal detection (warm, cold), pain thresholds (painful heat and cold), and pain to a defined suprathreshold painful stimulus will be determined (*50*). The Advanced Thermal Stimulator thermode (contact area of 16 x 16mm) was placed on the volar forearm. Testing starts at a neutral temperature (32°C) and increases (or decreases) with 1.5°C/s up to a maximum of 50°C (or down to a minimum of 0°C). First, thresholds of cold (CDT) and warm (WDT) detection were measured three times each, followed by an assessment of pain cold (CPT) and heat (HPT) thresholds. Volunteers were instructed to press a button when sensation changes to warm/cold or become painful; then, the thermode immediately returns to the baseline temperature (32°C). If the cut-off temperature (50°C or 0°C) was reached, the device automatically returns to the baseline temperature (32°C) to avoid tissue damage. The mean value from the three measurements was taken as the heat and cold pain threshold. In addition, the pain intensity to a suprathreshold heat pain stimulus was assessed by application of three test stimuli (45, 46, and 47°C, using a ramp of 8°C/s and time at the target of 7s). In addition, subjects will be asked about paradoxical heat sensations (PHS) during the thermal sensory limen (TSL) procedure of alternating warm and cold stimuli.

Modified von Frey hairs (Optihair2-Set; Marstock Nervtest, Schriesheim, Germany) that exert forces between 0.25 and 512 mN (geometric progression by factor 2) was used to assess the mechanical detection threshold (MDT). The end of the von Frey filaments was provided with a rounded tip (0.5 mm diameter) to avoid sharp edges that may activate nociceptors. Threshold determinations were made using an adaptive method of limits by a series of alternating ascending and descending stimulus intensities (up-and-down rule), yielding 5 just suprathreshold and 5 just subthreshold estimates. The geometric mean of these 10 determinations was represented the final threshold.

Standardized punctate probes performed the assessment of mechanical pain sensitivity and threshold with fixed intensities (8, 16, 32, 64, 128, 256, and 512 mN), and a contact area of 0·2 mm diameter will be used to determine mechanical pain sensitivity (MPS) and mechanical pain thresholds (MPT) (*49, 51*). Stimulators were applied at a rate of 2 s on, 2 s off in ascending order until the first percept of sharpness was reached to assess MPT. The final threshold was the geometric mean of five ascending and descending stimuli series. To determine MPS the same stimuli were applied in a pseudo-random order with a 10-sec interval in 5 runs; subjects were asked to give a pain rating for each stimulus on a ‘0–100’ numerical rating scale (’0’ indicating “no pain”, and ‘100’ indicating “most intense pain imaginable”). MPS was calculated as the geometric mean of all numerical ratings for each pinprick stimulus.

QST parameters were measured in their physical dimension and were weighted by transformation to the standard normal distribution (Z-transformation). Z-scores indicate gain (above “0”) or a loss (below “0”) of function across QST-parameters.

### Psychosocial baseline characterization by questionnaires

Prior incision injury, volunteers have completed a set of psychological questionnaires, including the Beck’s Depression Inventory (BDI), the state-trait anxiety inventory (STAI), the pain catastrophizing scale (PCS), and the pain sensitivity questionnaire (PSQ) to identify possible risk factors for incision-induced. The volunteer completed the questionnaires himself in a quiet neutral room in the presence of a male experimenter. The order of the questionnaires was randomized a priori by an Excel list for each volunteer individually.

PSQ is a self-rating instrument for the assessment of pain sensitivity, validated in healthy subjects and chronic pain patients (*52–56*)

Blood sampling and plasma processing

Blood samples (5ml) were collected by venipuncture of the contralateral arm under sterile conditions. Winged blood collection sets (BD Vactuainer® Safety-Lok™, needle gauge 21) were used to collect the blood in EDTA tubes (BD Vactuainer®, K_2_EDTA, 1·8 mg/ml). All tubes were gently rotated eight times by hand after sampling. Blood cells, including platelets, were removed from plasma by centrifugation at 2,000 x g for 15 minutes in a refrigerated centrifuge (4°C) to obtain blood plasma samples. The blood plasma was divided into 0.5 ml aliquots and stored at −20°C.

### Experimental incision

The experimental incision followed a protocol previously described(*23, 51, 57*). Briefly, the skin of one randomly chosen arm for incision was disinfected with 70% ethanol. An incision of 4 mm width and 7 mm depth was ‘applied’ with a sterile scalpel (No. 11), perforating the muscular fascia. A gauze swab stopped the bleeding of the wound with gentle pressing.

### Determination of incisional pain and hyperalgesic area

Subjects were asked to rate incision pain (IncP) on a numeric rating scale (NRS, 0-100) at time zero, every minute until 10 minutes, every five minutes until 1 hour, and finally 24 hours after incision. The area under the curve was determined for the first 60 minutes after incision, which represents acute incisional pain. Sensitivity to punctuate mechanical stimulation in the secondary zone (2), namely the secondary hyperalgesic area (HA), was determined using a conventional von Frey filament with 116 nN bending force. It was applied in eight imaginary lines at 45°, starting far distant from the putative hypersensitivity region centripetally directed towards the incision. All eight points were linked, transferred onto a paper, and the area was determined in Image J (https://imagej.nih.gov/ij/). HA determination was executed 1 h (HA1) post-surgically.

### Definition of responders for incision-induce outcome measures

Volunteers (V) were stratified into responder types depending on the IncP and the extent of the HA. The mean of the entire cohort was calculated for both outcome measures. In the following, volunteers with a larger IncP and larger hyperalgesic areas are designated as high-responders.

Being above or below the corresponding mean, volunteers were stratified into high (> mean) and low (< mean) responders assuming that the rating of post-operative pain reflects acute or the extent of hyperalgesia upon peripheral input and putative chronic pain vulnerability. Ultimately, we defined IncP and HA as reference parameters for psychophysical, psychometric, and proteomic analyzes to determine incision injury single marker and multi-feature prediction signatures.

### Sample preparation: protein digestion and sample clean-up

Plasma samples were neither fractionated nor depleted. Plasma samples were allowed to thaw on ice and sonicated in 15 sec intervals, depending on the degree of denatured components. 10 µL plasma, corresponding on average to 36,1 µg/µL were diluted in 5 mM TCEP, 1% SDC in 100 mM ABC (DR buffer). Samples were denatured and reduced for 1 hour at 60°C with agitation in a thermoshaker. 15 µL of the denatured mixture was further diluted with 100 mM ABC in a 1:1 ratio and alkylated with 10 mM IAA at room temperature for 30 minutes (kept in the dark). Quenching of samples was obtained by adding 10 mM DTT final for 15 minutes of incubation at room temperature. Samples were diluted with MS-grade H_2_O to perform protein hydrolysis in 50 mM ABC at pH 7,8 – 8,0. Tryp/rLys-C was added 1:50 (enzyme: protein) and incubated overnight (16 hours) at 37°C using a heated thermoshaker with heatable lid (40°C) to prevent condensation within the Eppendorf tube.

Removal of detergents was obtained by phase transfer following the protocol by Masuda and Ishihama (*58*). After discarding the surfactant-containing upper layer, the remaining peptides were desalted using solid-phase extraction (SPE). C_18_ MicroSpin (The Nest Group, USA) columns were utilized for SPE, adhering to the manufacturer’s instructions with the following modifications: bound peptides were subsequentially washed with 0.5% TFA and 0.2% TFA, two-step elution of peptides with 50 µL 50% ACN, 0.1% FA and 50 µL 80% ACN, 0.1% FA. Cleaned-up peptides were fully dried using a vacuum evaporator at 38°C.

### Liquid chromatography and mass spectrometry

Dried peptide samples were solubilized in MS buffer (1% ACN, 0.1% FA) and sonicated for 2 minutes. Peptide concentrations were determined using a UV/VIS Spectrometer at 280 nm/430 nm (IMPLEN, Germany). Autosampler vials contained 100 ng/µL sample and iRT (Biognosys, Switzerland) peptides for prediction of peptide retention times upon chromatographic separation (*59*). A pooled sample of V1-V3 (100 ng/µL) was run before each acquisition queue to control for consistent system performance.

NanoLC-MS/MS analyses were conducted on an Orbitrap Exploris 480 mass spectrometer coupled with an UltiMate 3000 UHPLC System (Thermo Scientific, Germany). In a direct setup, 2 µL peptides were separated on an Acclaim PepMap column (75 µm x 15 cm, C18, 2 µm, 100 Å) at a constant flow rate of 300 nl/min and injected to the mass spectrometer via a Nanospray Flex^TM^ ion source (Thermo Scientific, USA). The mobile phase consisted of [A] 0.1% FA, 1% ACN and [B] 0.1% FA, 100% ACN. MS acquisition included peptides eluting along a two-step linear gradient of 3% to 25% B in 60 minutes and 25% to 40% B in 20 minutes. The high organic phase (80% B, 10 minutes) and two subsequent short gradients (3% to 50% B 10 minutes; 80% B 4 minutes) within the same injection cycle were without MS acquisition to ensure minimisation of a column- and LC system-related carryover in a time-efficient manner. The column temperature was set to 40°C constantly. MS raw data were acquired in data-independent acquisition mode (DIA) in a scan range of 350 to 1154 m/z. MS1 spectra were recorded at a resolution of 120,000 and automatic gain control (AGC) target of 3e^6^ or 60 ms injection time, respectively. Corresponding MS2 scans were acquired at 30,000 resolutions with an AGC value of 3e^6^ and auto for injection time. Each DIA segment contained 15 spectra of 18 Da windows with a stepped collision energy of 25, 27, and 30. A total of three full scans each followed by symmetrically segmented isolation windows was needed to cover the entire MS scan range. All spectra were recorded in profile mode.

### Quantitative LC-MS data analysis

The raw DIA data files were processed with the open source program DIA-NN 1.8.1 (*60*). As spectral library, an in-house library stemming from human plasma depletion experiments was used. The spectral library contained 4590 protein isoforms, 2889 protein groups and 17809 precursors in 14923 elution groups. For the *in vitro* digestion, Trypsin/P was chosen and a maximum of three missed cleavages. Peptide length was set to 5 to 52 amino acids. Cysteine carbamidomethylation was chosen as a fixed modification and as possible variable modifications N-term methionine excision, methionine oxidation, and/or N-term acetylation were set. The maximum number of variable modifications allowed was 3. RT-dependent cross-run normalization and Robust LC (high accuracy) options were selected for quantification. The R package, DiaNN (https://github.com/vdemichev/diann-rpackage; (*60*)) was used to extract the MaxLFQ (*61*) quantitative intensity of gene groups for all identified protein groups with q-value < 0.01 as criteria at precursor and gene group levels. Due to poor data quality, V7 was excluded from all analyses. All raw data files have been uploaded to the Proteome Xchange Consortium (*66*) via the PRIDE partner repository with the dataset identifier PXD033592.

### Network-based drug repositioning with PharmOmics

The network-based repositioning tool within the open-source application PharmOmics (http://mergeomics.research.idre.ucla.edu/runpharmomics.php) (*62*) was used. As input genes, the high responder prognostic markers were taken. The analysis was run for each of the outcome measures (IncP, HA1) separately. The signature type was set to “Meta”, species to “Human” and as the background network we chose (i) “Sample Multi-tissue Network” (provided by PharmOmics) and (ii) a protein-protein interaction network created with STRING (https://string-db.org/ (*67*), analysis performed in January 2024) based on our own identified plasma proteins (if more than 1 protein name was given, the first entry was kept; duplicates were deleted yielding a network with 285 nodes). The output was filtered for the tissues “hematopoietic system” and “immune system”.

### Statistical analyzes, bioinformatic processing, and post-hoc analyzes

Bioinformatic analysis of LC-MS data was mainly performed in Spectronaut 15 (Biognosys, Switzerland) or in the freely available Perseus software (https://maxquant.net/perseus/). Feature-selection of prognostic marker and marker combinations was performed in the statistical environment R. If not reported otherwise, data were analyzed using OriginLab PRO.

#### Differential abundance analysis

Mean log_2_ ratios (high/low) were calculated in Spectronaut 15 for each protein ID, and unpaired t-testing was performed. To control the false discovery rate (FDR), obtained p-values were Benjamini-Hochberg (BH)-adjusted and proteins with qvalue < 0.05 are considered to be significantly different.

#### Network analysis

Cytoscape (version 3.8.2, available at cytoscape.org28) was used to identify species-specific and pain phenotype-specific protein networks (Term-term-interactions TTI-networks). Functional grouping within these networks was achieved using AutoAnnotate version 1.3. Analysis revealed significantly enriched GO terms (P-values ≤ 0.05) within these functionally grouped networks, effectively reflecting the relationships between these terms.

#### Logistic regression analyses

This study aimed to propose prediction models for the defined outcomes IncP, and HA1. Therefore, we employed logistic regression models in R to identify combinations of available candidates (i.e. covariates) that correlate with a high probability for high-responders. We developed an analysis pipeline comprising the following steps: (I) data preprocessing, (II) model building, and (III) evaluation of model robustness. Data preprocessing applies normalization per candidate and a linear shift to a beneficial data range for subsequent analysis. This data shift is introduced only for technical reasons and alters data in any relevant manner. Then, the normalized and shifted predictors are analyzed using the *combiroc* package(*63*). A wrapper function has been programmed which allows to firstly analyze each candidate variable individually, filter predictors for a selected minimal AUC value and subsequently evaluate all combinations between all selected candidates. All models are fitted according to the *comibroc* default procedure, i.e. to fit logistic regression models. All combinations generated in step II are evaluated by leave-one-out cross-validation (LOOCV). We defined deviance measures by calculating the mean squared difference between the AUC from all volunteers and the individual LOOCV AUCs. We used the overall AUC and its deviance to rank each predictor or combination. The selection of the optimal predictor/combination is a multi-criteria optimization problem (maximize AUC & minimize deviance). Thus, more than one combination could be optimal in the sense of a Pareto front. In this case, an expert decision has been applied based on other criteria, e.g. applicability or feasibility of the specific measures in the clinical context. Our pipeline also allows restricting the search space to pre-specified subsets of combinations - e.g., we can restrict the algorithm to search within all combinations of maximal length three and pre-specify that each candidate shall be of a distinct dimension (e.g. 1x psychophysical parameter + 1x psychological parameter + 1x blood plasma protein). The receiver operator curves (ROC) of the fitted logistic regression models yield area under the curve (AUC) values.

## Data Availability

All data produced in the present study are available upon reasonable request to the authors

## Acknowledgments

AI-powered applications, namely ProWritingAid (ProWritingAid Inc.; Canada) and ChatGPT (OpenAI, United States), were employed for linguistic processing objectives, encompassing grammar correction and the enhancement of word selection.

## Funding

Deutsche Forschungsgemeinschaft (DFG) (SCHM 2533/6e1 and SCHM 2533/4e1 to MS, PO1319/3e1 to EPZ). Federal Ministry of Education and Research (BMBF), Germany, to EPZ (01KC1903). University of Vienna to MS.

## Author contributions

Study design: DS, MS, EPZ

Experiments: DS, JRS, CK, MvdB, BP, DG, MF, PKZ, MS, EPZ

Data analysis: DS, JV, JRS, CK, DG, MS

Writing first manuscript draft: DS, JRS, CK, MS, EPZ Figure preparation: DS, MS, EPZ, JRS, JV

Revising manuscript: MvdB, CK, DG, MF, PKZ, MS, EPZ

## Competing interests

MS received research awards and travel support by the German Pain Society (DGSS) both of which were sponsored by Astellas Pharma GmbH (Germany). MS received one-time consulting honoraria by Grunenthal GmbH (Germany). None of these funding sources influenced the content of this study, and MS declares no conflict of interest. During the past 5 yr, EPZ received financial support from Grunenthal for research activities and from Grunenthal, Novartis (Switzerland), Medtronic for advisory board activities, lecture fees, or both. None of this research support/funds was used for or influenced this manuscript, and EPZ declares no conflict of interest. The remaining authors declare that they have no conflicts of interest.

## Data and materials availability

All data are available in the main text or the supplementary materials. All protein raw data files have been uploaded to the Proteome Xchange Consortium (*66*) via the PRIDE partner repository with the dataset identifier PXD033592.

## Notes

### Clinical Trial

DRKS00016641

### Author Declarations

University Hospital Muenster of the local Ethics Committee of the Medical Faculty (registration no 2018-081-b-S)

