## Supplementary Material for "BLOOD PROTEOMICS AND PAIN - A TRANSLATIONAL STUDY TO PROGNOSTICATE PAIN PHENOTYPES AND ASSESS NEW BIOMARKERS FOR PREVENTING PAIN IN HUMANS"

**Table of supplementary contents**


| Supplementary tables |
| --- |
| **Table S1** |
| **Table S2** |
| **Table S3** |

| Supplementary figures | |
| --- | --- |
| Fig. S1 | Details of protein groups quantified per sample and top scoring pathways and GO annotation |
| Fig. S2 | Post-incisional syptoms |

**Supplement Table 1 In- and exclusion criteria**

| Criteria | Inclusion | Exclusion |
|---|---|---|
| **Gender** | Male | Female |
| **Age** | 18-40 y | > 40 y |
| **Health Status** | Healthy | Pre-existing diseases<br>• Diabetes<br>• cardiac diseases<br>• pruritus<br>• skin diseases (arm)<br>• neurological diseases |
| **Treatment** | No chronic use of medication | Regular analgesic treatment<br>(< 3days before experiment) |
| **QST** | Unobtrusive | Obtrusive one day before incision |

### Supplement Table 2. Baseline QST raw values of volunteers

| QST | Control Area | Test Area |
|---|---|---|
| **CDT, °C** | -1.32 ± 0.53 | -1.53 ± 0.92 |
| **WDT, °C** | 2 ± 0.86 | 2.2 ± 0.69 |
| **TSL, °C** | 0.6 ± 0.9 | 0.8 ± 0.8 |
| **CPT, °C** | -13.38 ± 7.6 | -11.83 ± 8.49 |
| **HPT, °C** | 9.88 ± 3.9 | 10.20 ± 3.43 |
| **MDT, mN** | 1.33 ± 1.53 | 1.23 ± 1.36 |
| **MPT, mN** | 55.82 ± 32.11 | 74.4 ± 142.88 |
| **WUR** | 3.13 ± 3.1 | 3.1 ± 3.11 |
| **VDT, /8** | 7.02 ± 0.38 | 7.11 ± 0.5 |
| **PPT, kg** | 4.45 ± 1.16 | 4.67 ± 1.16 |

Data are expressed as mean ± SD. Bold numbers indicates *P < 0.05, **P < 0.01, or ***P < 0.001 versus Control Area. Comparisons between areas (control area vs test area) were performed using paired t-test. Abbreviations: CDT, cold detection threshold; CPT, cold pain threshold; HPT, heat pain threshold; MDT, mechanical detection threshold; MPT, mechanical pain threshold; PPT, pressure pain threshold; QST, quantitative sensory testing; TSL, thermal sensory limen; VDT, vibration detection threshold; WDT, warmth detection threshold; WUR, wind-up ratio.

**Supplement Table 3. Pyschosocial questionnaires**

| Questionnaire | | Score Range | Median [95%CI] | Results |
|---|---|---|---|---|
| **Beck-Depressions-Inventar (BDI-II)** **0-9 clinically unremarkable** **10-18 mild** **19-29 moderate** **>29 severe** | | 0-45 (a lower score denoting a better outcome) | 4 [4 to 7.3] | Clinically unremarkable |
| **Revised Life Orientation Test (LOT-R)** | Optimism | 0-12 (a higher score denoting a better outcome) | 10 [8.2 to 10.1] | Clinically unremarkable |
| | Pessimism | 0-12 (a higher score denoting a better outcome) | 9 [7.8 to 9.5] | Clinically unremarkable |
| | Total | 0-24 (a higher score denoting a better outcome) | 19 [16.2 to 19.3] | Clinically unremarkable |
| **Pain Catastrophizing Scale (PSC)** **>30 severe** | | 0-52 (a lower score denoting a better outcome) | 12 [9.1 to 14.9] | Clinically unremarkable |
| **Pain Sensitivity Questionnaire (PSQ)** | Minior | 0-10 (a lower score denoting a better outcome) | 2.2 [1.8 to 2.4] | Clinically unremarkable |
| | Moderate | 0-10 (a lower score denoting a better outcome) | 3.9 [3.3 to 4.4] | Clinically unremarkable |
| | Total | 0-10 (a lower score denoting a better outcome) | 3.1 [2.6 to 3.4] | Clinically unremarkable |
| **State-Trait Anxiety Inventory (STAI)** | X1 | 20-80 (a lower score denoting a better outcome) | 34 [31.3 to 36.1] | Clinically unremarkable |
| | X2 | 20-80 (a lower score denoting a better outcome) | 35 [33.4 to 39.6] | Clinically unremarkable |

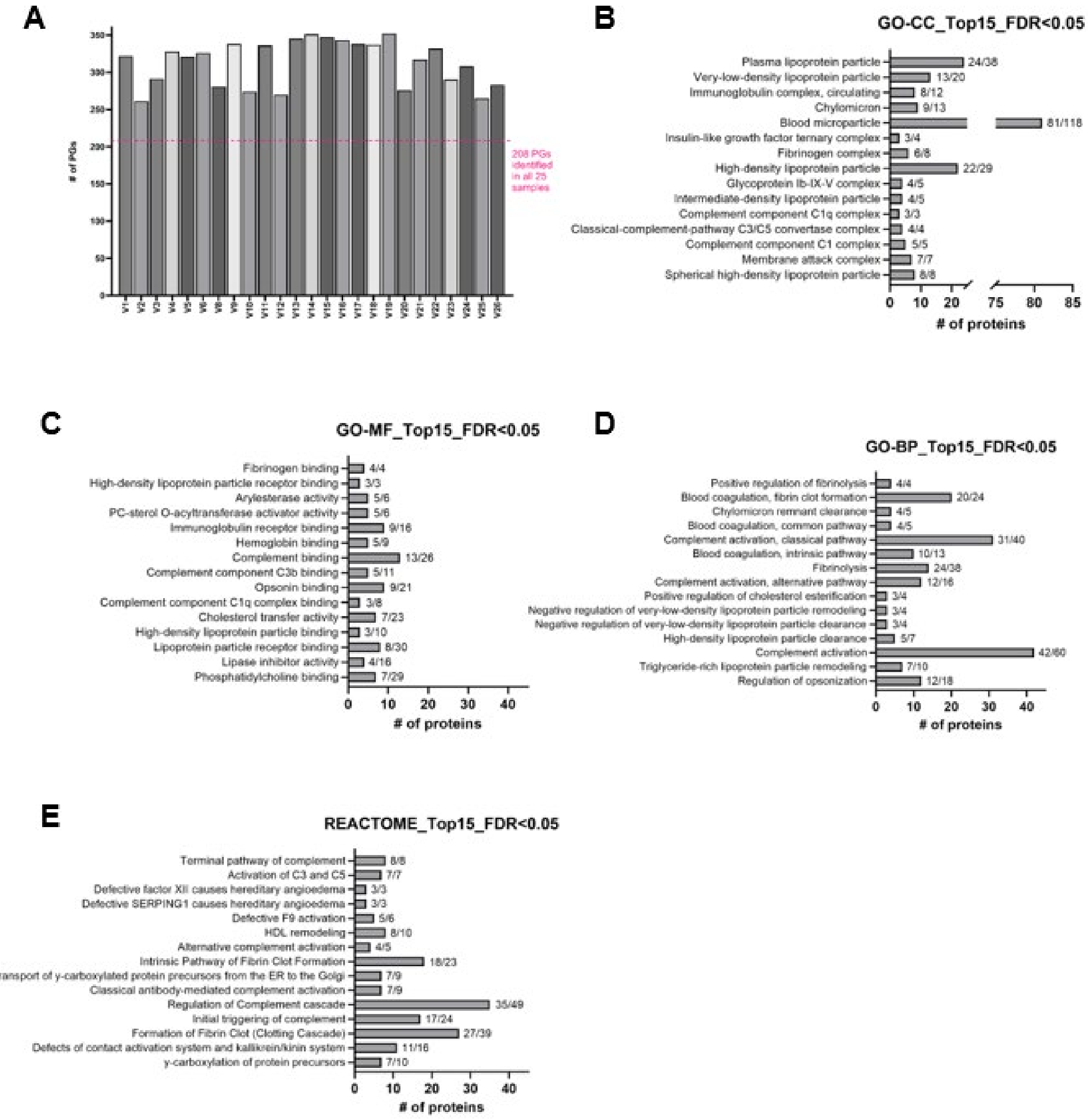


**Supplementary Figure 1.**: Details of protein groups quantified per sample and top scoring pathways and GO annotation
(A) Bar chart shows the number of protein groups (PGs) identified and quantified across all 25 volunteers. Dashed line in red denotes the 208 PGs that were quantified in all of the samples.
(B-E) Top 15 GO-CC (B), MF (C), BP (D) terms, and REACTOME (E) pathways ranked by their coverage. Protein groups were submitted to GO annotation and REACTOME pathway analysis via the STRING web interface (see methods for details, full lists in suppl table X). Only pathways with an FDR <0.05 and at least 3 proteins included and identified were considered. Numbers next to column show observed proteins/all proteins included in pathway and/or term).

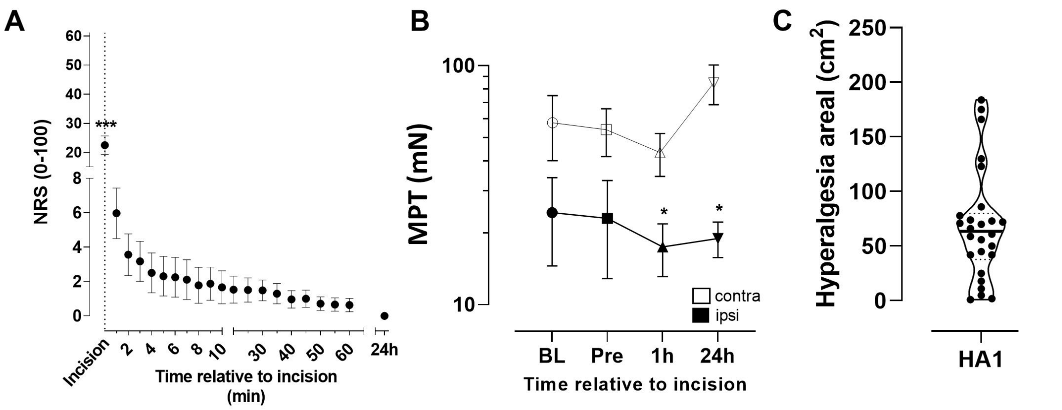


**Supplementary Figure 2. Post-incision syptoms** (A) Pain related to the incision was rated using the numeric rating scale (NRS, 0;100). Pain was maximal during the incision and decreased considerably within the first ten minutes (1 measurement per minute). Up to 1 h, additional NRS determinations in 5-minute steps exhibited a weaker slope. Pain was completely absent after 24 h. Sixty minutes after the incision, integration of the area under the curve (AUC) of all values resulted in volunteer specific $NRS_{AUC}$ values. N=26, Mean with SEM, Holm-Sidak multiple comparison test, ***P<0.001.

(B) Mechanical pain thresholds (MPT) 24 h before the incision (baseline, BL), pre-incision (Pre), and 1 h or 24 h post-incision. MPT was determined in milli Newton (mN) at the contra- (white) and ipsi-lateral (black) side. Upon incision, i.e. on the ipsilateral side, MPT was significantly decreased 1 h and 24 h post-surgically. N=26, Mean with SEM, Holm-Sidak multiple comparison test, *P<0.05.

(C) Determination of the secondary hyperalgesic area (HA) in $cm^2$ to mechanical stimuli (von Frey filament) in 26 volunteers 1h post-incision. N=26, Median, 25th and 75th quartils (dashed line) truncated violin plot.
